# A gene-diet interaction-based score predicts response to dietary fat in the Women’s Health Initiative

**DOI:** 10.1101/19004937

**Authors:** Kenneth Westerman, Qing Liu, Simin Liu, Laurence D. Parnell, Paola Sebastiani, Paul Jacques, Dawn L. DeMeo, José M. Ordovás

**Affiliations:** JM-USDA Human Nutrition Research Center on Aging, Boston, MA; Department of Epidemiology, Brown University School of Public Health, Providence, RI; Department of Biostatistics, Boston University School of Public Health, Boston, MA; Channing Division of Network Medicine, Department of Medicine, Brigham and Women’s Hospital, Boston, MA; IMDEA Alimentación, CEI, UAM, Madrid, Spain; Centro Nacional de Investigaciones Cardiovasculares (CNIC), Madrid, Spain

## Abstract

While diet response prediction for cardiometabolic risk factors (CRFs) has been demonstrated using single SNPs and main-effect genetic risk scores, little investigation has gone into the development of genome-wide diet response scores. We sought to leverage the multi-study setup of the Women’s Health Initiative cohort to generate and test genetic scores for the response of six CRFs (body mass index, systolic blood pressure, LDL-cholesterol, HDL-cholesterol, triglycerides, and fasting glucose) to dietary fat. A genome-wide interaction study was undertaken for each CRF in women (n ∼ 10000) not participating in the Dietary Modification (DM) trial, which focused on the reduction of dietary fat. Genetic scores based on these analyses were developed using a pruning-and-thresholding approach and tested for the prediction of one-year CRF changes as well as long-term chronic disease development in DM trial participants (n ∼ 5000). One of these genetic scores, for LDL-cholesterol (LDL-C), predicted changes in the associated CRF. This 1760-variant score explained 3.4% of the variance in one-year LDL-C changes in the intervention arm, but was unassociated with changes in the control arm. In contrast, a main-effect genetic risk score for LDL-C was not useful for predicting dietary fat response. Further investigation of this score with respect to downstream disease outcomes revealed suggestive differential associations across DM trial arms, especially with respect to coronary heart disease and stroke subtypes. These results lay the foundation for the combination of many genome-wide gene-diet interactions for diet response prediction while highlighting the need for further research and larger samples in order to achieve robust biomarkers for use in personalized nutrition.

## Introduction

Nutrigenetics approaches, in which genetic information is used to predict response to dietary inputs, are central to the emerging promise of personalized nutrition for cardiometabolic risk reduction. Inter-individual differences in food preferences, metabolism, detoxification, excretion, etc. affect our responses to diet, in a similar manner to the well-studied field of pharmacogenomics (1). Ideally, genotype-based nutrigenetic investigations would be conducted in large-scale dietary interventions. Two notable examples are the PREDIMED and POUNDS LOST trial, with significant findings including the interaction of a *TCF7L2* variant with a Mediterranean diet pattern for glycemic traits (2) and the interaction of a *PCSK9* variant with dietary carbohydrate for insulin resistance (3). However, such intervention-based studies are able to examine only a single dietary change (whether food, nutrient, or pattern) at a time, and are often limited to lower sample sizes (4).

To allow for more flexibility and greater sample sizes, gene-diet interactions (GDIs) are more commonly investigated in observational datasets. There is a rich literature of GDI discovery in the cardiometabolic realm. Typically, these focus on cardiometabolic risk factors in relation to biology-based candidate genes/variants (5,6), but some have looked at clinical outcomes (e.g. MI (7)). Other approaches use main-effect genetic risk scores, such as that for obesity interacting with sugar-sweetened beverage intake to influence anthropometric traits (8,9).

Characterization of individuals based on single or small groups of single nucleotide polymorphisms (SNPs) likely neglects important signal elsewhere in the genome, especially when dealing with highly polygenic cardiometabolic traits. Thus, for effective personalized nutrition approaches to be realized, it is necessary to integrate signals across the genome. A few investigations have explored GDIs genome-wide, such as for dairy and BMI (10) and for various dietary components and colorectal cancer (11). However, genome-wide interaction studies (GWIS) can be problematic due to the lower statistical power inherent in gene-environment interaction analyses (12). Furthermore, the potential for confounding and reverse causation (i.e. cardiometabolic risk impacting dietary behavior) in statistical interactions from observational data means that GDIs may not always predict modification of the risk factor in question after a dietary intervention.

In order to provide proof-of-concept for the use of GDIs in developing comprehensive diet response genetic scores, we sought to develop a genome-wide, GDI-based dietary fat response score for each of a series of cardiometabolic risk factors (CRFs). We performed genome-wide interaction studies for six CRFs, prioritizing sites with documented nominal main effects in prior genome-wide association studies, and used these intermediate results to derive fat response scores (FRS) for each CRF. We tested the performance of these scores in the fat reduction-focused Women’s Health Initiative Dietary Modification trial, finding that an FRS for LDL-cholesterol (LDL-C) predicts 1-year LDL-C changes selectively in the intervention arm. Furthermore, we found associations of the LDL-C FRS with incident coronary heart disease and stroke subtypes specifically in the fat-reduction arm of the trial over approximately 22 years of follow-up.

## Methods

### Women’s Health Initiative Dataset

The Women’s Health Initiative study consists of a series of substudies: three clinical trials (related to cancer, cardiovascular disease, and osteoporosis) and an observational study (13). Over 160,000 participants were enrolled between 1993-1998, with the ability to enroll in up to 3 of the clinical trials simultaneously. For the purposes of this analysis, participants were categorized based only on whether or not they were enrolled in the dietary modification (DM) trial, which randomized almost 50,000 women to a low-fat diet or a control diet with no recommended dietary changes, with primary outcomes being incidence of breast and colorectal cancers and heart disease [(14). Study of these participants conformed to the ethical guidelines outlined in the Declaration of Helsinki, and this research was approved by the Tufts Health Sciences IRB (protocol 12592).

Participants were comprehensively screened at baseline, including physical measurements, blood sample collection, and questionnaire administration, while only a subset of participants provided blood samples or returned questionnaires during later visits. The food frequency questionnaire (FFQ) was designed specifically for the WHI study, emphasizing specific foods and preparation methods to maximize its sensitivity to changes in fat intake (15).

Phenotype data were accessed from dbGaP (accession: phs000746.v2.p3). Values shown in Table 1 only pertain to women whose genotypes were measured in one of a series of follow-up studies. For gene-diet interaction analyses, SBP, LDL-C, and GLU were adjusted for medication use: LDL-C and GLU values were divided by 0.75 for those on lipid-lowering and anti-diabetic medication, respectively, and SBP values were increased by 15 mmHg for those on anti-hypertensive medication. This type of adjustment for medication use has precedent in gene-environment interaction analyses (16). Cardiovascular risk factors (CRFs) were winsorized at 5 standard deviations from the mean and those other than LDL-C (BMI, SBP, HDL-C, TG, and GLU) were log-transformed prior to analysis. Longitudinal risk factor changes were calculated in DM trial participants as the difference between baseline and year 1. Adjudicated time-to-event data for chronic disease outcomes (coronary heart disease, myocardial infarction, ischemic stroke, hemorrhagic stroke, and non-CVD death) were collected, while diabetes incidence was defined as the self-report of any of: diabetes pills, insulin treatment, or general treatment for diabetes. Follow-up data was available for approximately 22 years following enrollment. Phenotype data processing was performed using R version 3.4.3 (17) and Python version 3.6.0.

**Table 1:** Baseline characteristics of European-ancestry participants in the Women’s Health Initiative Study (n = 17304 in total)

|  | DM trial | Non-DM trial |
| --- | --- | --- |
| Sample size | 6173 | 11131 |
| Age | 66 (61-70) | 68 (64-72) |
| Current smoking | 403 (7%) | 994 (9%) |
| Lipid-lowering medication | 655 (11%) | 1454 (13%) |
| Hypertension medication | 2334 (38%) | 4023 (36%) |
| Diabetes medication | 300 (5%) | 560 (5%) |
| Body mass index (BMI; kg/m<sup>2</sup>) | 28.7 (25.2-32.8) | 26.9 (23.7-30.9) |
| Systolic blood pressure (SBP; mm Hg) | 128.5 (117.5-140) | 144 (132-156) |
| LDL cholesterol (LDL-C; mg/dL) | 150.8 (127.5-175) | 161 (134-191) |
| HDL cholesterol (HDL-C; mg/dL) | 50 (42.5-58) | 51 (44-60) |
| Triglycerides (TG; mg/dL) | 137 (100-194) | 128 (92-179.6) |
| Fasting glucose (FG; mg/dL) | 97 (90-107) | 97 (90-113.3) |

**Table 2:** Responder score effects on 1-year CRF changes in DM trial participants

| Risk factor | # SNPs in risk score | N | Std. effect size | P-value | Std. effect size | P-value |
| --- | --- | --- | --- | --- | --- | --- |
| BMI | 6042 | 1988 | 0.03 | 0.221 | 0.02 | 0.253 |
| SBP | 1536 | 2004 | 0.03 | 0.196 | 0.04 | 0.061 |
| LDL-C | 1760 | 145 | -0.18 | 0.026 | -0.19 | 0.007 |
| HDL-C | 1731 | 150 | -0.06 | 0.471 | -0.06 | 0.483 |
| TG | 1774 | 150 | -0.14 | 0.066 | -0.14 | 0.064 |
| FG | 1924 | 281 | 0.02 | 0.791 | 0.03 | 0.570 |

### Genotype data and preprocessing

Imputed genotype data were retrieved from dbGaP (accession: phs000746.v2.p3) as a harmonized set of imputation outputs from a series of genotyping studies involving WHI participants. Prior to imputation, study-specific quality control steps had been undertaken on directly-genotyped SNPs, with filters based on sample and call rate, Hardy-Weinberg equilibrium, and minor allele frequency. Phasing had been performed for autosomes using BEAGLE, followed by imputation using minimac (MACH for the SHARe study subset). After download from dbGaP, variants were converted from dose format using dose2plink (http://genepi.qimr.edu.au/staff/sarahMe/dose2plink), filtered for imputation R-squared > 0.3 and minor allele frequency (MAF) >0.001, and annotated with rsIDs, loci, and allelic information using the 1000 Genomes Phase 3 download from dbSNP (download date: April 13, 2018). Only variants passing the imputation quality threshold in all genotyping sub-studies were included in the final dosage dataset. For score development and calculation, imputed variant dosages were converted to hard-calls and set to missing if the estimated dosage was not within 0.1 of an integer allele count. Post-imputation genotype data processing was performed using PLINK 2.0, while clumping and score calculation were performed using PLINK 1.9 (18).

### Genome-wide interaction studies

A genome-wide interaction study was performed for each of the six cardiometabolic risk factors. The genome-wide scan used an additive genotype model, adjusted for fixed effects including dietary fat (binary: % of kcals above or below the median), total kcals per day, age, five ancestry principal components, and genotyping sub-study. Genotyping was performed in a series of ancillary studies in WHI including Hip Fracture, GARNET, WHIMS+, GECCO (initial or CytoSNP), and AS264/MOPMAP. (Many participants were also genotyped as a part of the SHARe effort, but those women were of African American and Hispanic ancestry and thus were not included in the GWIS portion of this study.) Five of six sub-studies were adjusted for as binary variables to avoid collinearity. The primary estimand of interest was the interaction term between dietary fat and minor allele count at the SNP of interest. Interaction analyses were carried out using PLINK 2.0 (18). Variants of interest were annotated to genes using Annovar (19).

Gene-environment interaction power calculations for single SNPs were performed using the Quanto tool (20). The following assumptions were made: additive model; variance explained by genotype alone = 0.5%; and binary environment with 50% prevalence and explaining 10% of variance. (Note: there is no effect of minor allele frequency in this case given that variances explained are directly specified.)

### Genetic responder score construction and evaluation

To prioritize variants for inclusion in genetic responder scores, nominal (p < 0.05) variants for main-effect on each risk factor were retrieved from large-scale meta-analyses: GIANT for BMI (21); International Consortium for Blood Pressure for SBP (22); Global Lipid Genetics Consortium (GLGC) for LDL-C, HDL-C, and TG (23); and MAGIC for fasting glucose (24). Each FRS was constructed using summary statistics for the diet-SNP interaction terms from the associated GWIS. SNPs were filtered for nominal main-effect relationships using the above meta-analyses, and interaction summary statistics were used as input to a pruning-and-thresholding (P&T) procedure (using the “–clump” function in PLINK 1.9), with a seed threshold of p=0.05 and an LD threshold of r^2^=0.5. The linkage disequilibrium references for the procedure was calculated from genotypes of the white DM trial participants. Genetic fat response scores (FRS) for each individual were then calculated as the weighted sum of allelic dosages for variants selected by the P&T procedure, with weights corresponding to the GWIS interaction term estimates. This type of diet interaction-based genetic score development has been described previously, for example in a Korean cohort with respect to body fat changes (25). A genetic risk score for LDL-C was created using the GLGC LDL-C meta-analysis summary statistics and the same P&T method and parameters as was used for the interaction analyses, resulting in a 26467-SNP score.

FRS were used to test for discrimination of changes in CRFs over the first year of the DM trial. Risk factor changes were assessed using linear models in participants in the intervention arm, with and without adjustment for baseline CRF levels. As a sensitivity analysis, p-values were calculated in separate models for interaction of the genetic score with 1) trial arm (control vs. dietary modification), and 2) observed fat reduction (negative vs. positive 3-year change in FFQ-estimated dietary fat). GRS were further tested for prediction of chronic disease development during follow-up across DM trial strata. Time-to-event for each of coronary heart disease, myocardial infarction, ischemic stroke, hemorrhagic stroke, and non-CVD death were used to fit age-adjusted Cox proportional hazards models, including a random effect term for genotyping sub-study (*cluster()* term in the *coxph* function call). Estimated log-hazard ratios were extracted from regressions conducted in the following strata: 1) DM trial intervention arm, 2) DM trial control arm, 3) DM trial intervention arm filtered for participants with 1-year fat reduction based on FFQ, and 4) DM trial control arm filtered for participants with 1-year fat increase based on FFQ.

## Results

### Dietary fat responder score development

A series of genome-wide interaction studies (GWIS) were undertaken in cross-sectional data from the Women’s Health Initiative. These GWIS incorporated only women who did not participate in the dietary modification (DM) trial, using imputed genotypes along with baseline self-reported dietary intakes (from food frequency questionnaires) and fasting blood biomarkers. Baseline characteristics of these women, along with those participating in the DM trial, are shown in Table 1.

Preliminary power calculations were undertaken, based on parameter assumptions including a modest SNP main effect (0.5%) under an additive model and a binary environment with 50% prevalence explaining 10% of the outcome phenotypic variance. The results showed that, at the sample sizes available for European ancestry non-DM participants (7000-10000 individuals for each cardiometabolic risk factor (CRF)), this analysis was powered to detect only moderately large interaction effects (GxE variance explained greater than approximately 0.5%) at genome-wide significance (Supp. Table S1).

Dietary fat response scores were generated for each CRF using results from the corresponding GWIS analysis. Linear regression models were fit for log-transformed baseline CRF values (other than LDL-C), incorporating gene-dietary fat interactions (dietary fat represented as a binary % of total calories above or below the median) while adjusting for age, total calories, five genetic principal components, and genotyping sub-study. Q-Q plots of the GWIS results showed that genomic inflation was fairly well-controlled (Supp. Figure S1). For each CRF, the associated summary statistics (corresponding to the fat-genotype interaction term estimates) were filtered to include only those with nominal main-effect associations in large-scale published GWAS. This filter was informed by the power analysis above and chosen as a compromise between discovery and statistical power (alternative results using either a more stringent threshold or no filtering are shown in Supp. Table S2). A pruning-and-thresholding method was used to generate six FRS from these individual sets of summary statistics along with genotypes from the WHI DM participants as a linkage disequilibrium (LD) reference. Using parameters of seed p-value=0.05 and LD r^2^<0.5, six sets of score weights were generated, with relevant SNP set sizes ranging from 1536 (SBP) to 6042 (BMI). Scores were then calculated as the weighted sum of allele counts across SNPs, normalized by the number of non-missing SNPs per individual.

### Dietary fat responder score assessment

As the scores were developed to predict a positive interaction with dietary fat intake, the expected direction of the FRS effect on risk factors in the present fat-reduction trial would be negative. Of the fat response scores examined, only the LDL-C fat response score (LDL-FRS) was predictive at p < 0.05 of the associated CRF change in DM trial participants in the fat-reduction arm (passing a Bonferroni correction for the 6 CRFs tested in baseline-adjusted sensitivity models). For this score, the standardized effect size was −0.18 (corresponding to a 5.1 mg/dL greater decrease in LDL-C per score standard deviation; p=0.026). We note that the sample size of European-ancestry DM trial participants with follow-up measurements was much smaller for biochemical variables (n∼150) compared to BMI and SBP (n∼2000). Using the score developed in European-ancestry individuals, score performance was then tested in a combined-ancestry group including Black and Hispanic individuals, which almost doubled the sample size (Supp. Table S3). While some traits showed strong relationships (e.g. SBP), the signs of many were in a counter-intuitive direction, including a flip in sign for the previously-strong LDL-C relationship, suggesting that these results reflect primarily differences in ancestry rather than the intended biological differences. This observation was reinforced by the lack of association of the score with CRF changes in either Blacks or Hispanics alone.

Based on its observed association in European-ancestry participants, the LDL-FRS was investigated further. Linear models showed that the LDL-FRS accounted for 3.4% of the variance in 1-year LDL-C changes in the DM intervention arm. In baseline-adjusted models, this figure rose slightly to 3.9, based on the R^2^ change from a baseline-only model. Additional models confirmed an interaction between DM trial arm and the LDL-FRS (p=0.011), supporting the specificity of this score for the fat-reduction arm. The LDL-FRS also showed specificity for LDL-C in that it did not predict changes in any other CRF (Supp. Table S4). The 1760 component SNPs were annotated using Annovar (19), revealing a predominance of intergenic and intronic variants and a set of genes with high numbers of independent SNPs contributing to the score (Figure 1a-d). Top genes by number of contributing SNPs included *CSMD1, PTPRD*, and *RGS12*.

**Figure 1:**
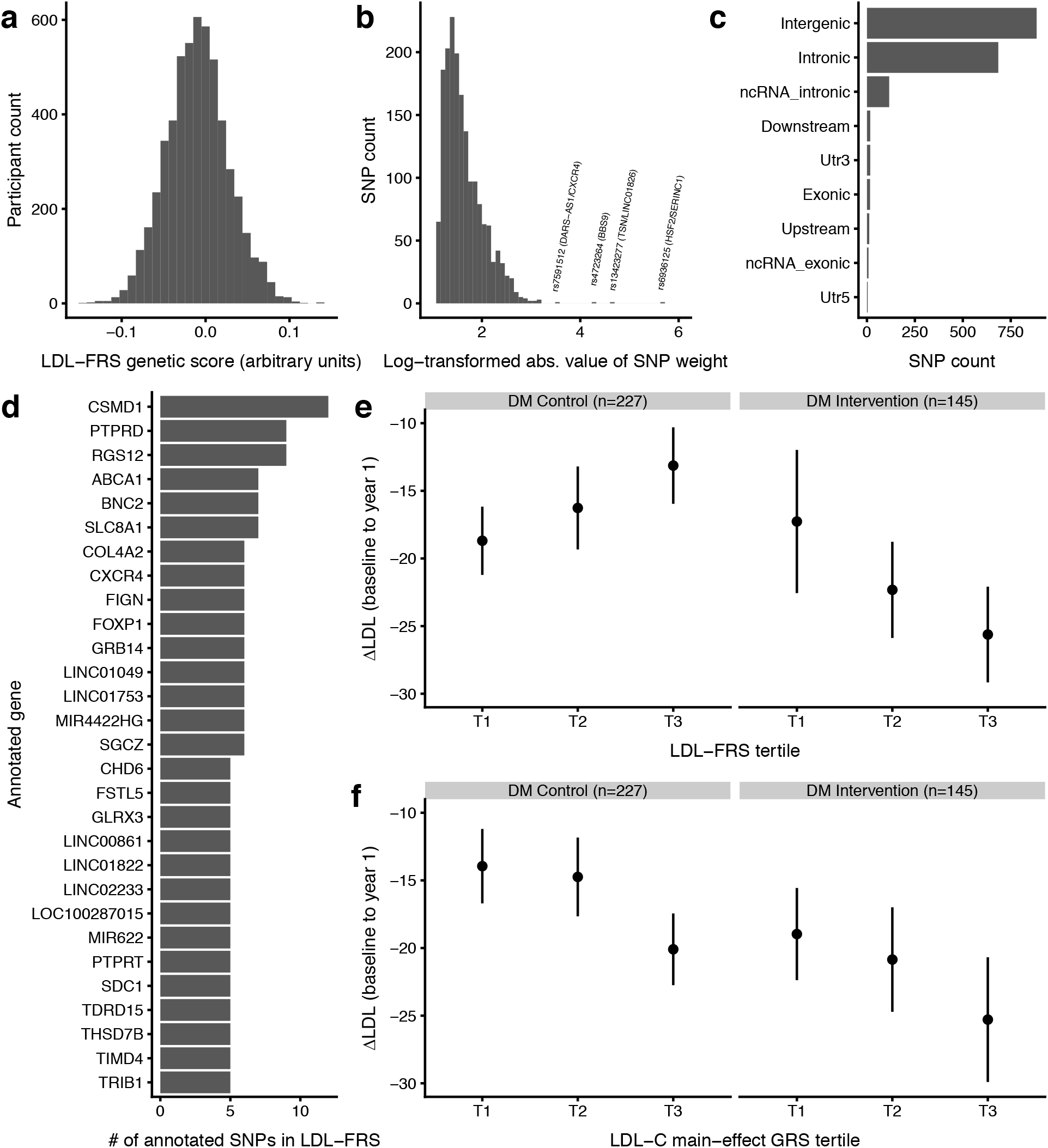
LDL-FRS characterization. a) Distribution of LDL-FRS in WHI DM trial participants. b) Distribution of SNP weights constituting the LDL-FRS (shown as the natural log-transformed absolute values of the true weights). c) SNP counts in different loci types for LDL-FRS constituent SNPs. d) Genes are summarized by the number of annotated SNPs in the LDL-FRS (genes with at least 5 component SNPs are shown). e-f) 1-year changes in LDL-C in DM trial participants as a function of genetic scores. Mean changes in LDL-C (y-axis) are shown as a function of either LDL-FRS (e) or LDL-GRS (f) tertile (x-axis). Error bars represent standard errors. LDL-FRS: LDL-C fat response genetic score, LDL-GRS: LDL-C main-effect genetic score.

Differences in mean LDL-C changes during the DM trial across genetic score strata are shown in Figure 1e,f. As suggested by the regression results, those in the control arm trended towards less strong LDL-C reductions in higher LDL-FRS strata, while those in the fat-reduction arm showed the opposite trend. Furthermore, isolation of individuals at the highest extreme of the score (top 10%) revealed an LDL-C reduction of almost double that of the rest of the DM intervention group (−39.0 versus −20.0 mg/dL). For comparison to the FRS, a main-effect genetic risk score (GRS) for LDL-C was developed using summary statistics from the Global Lipid Genetics Consortium meta-analysis (23) and an identical pruning-and-thresholding procedure to that used for the GDI-based scores. As expected, this score was strongly predictive of baseline LDL-C concentrations (p=4.53e-27). However, unlike the GDI-based score, the GRS did not predict LDL-C changes in the DM intervention group (p=0.15; stratum-specific mean changes in Figure 1f).

### LDL-FRS association with chronic disease outcomes

Next, the LDL-FRS was tested for relationships with incident disease outcomes over approximately 22 years of follow-up (Figure 2). Cox proportional hazards models were used, adjusting for age at baseline and including a random effect for genotyping sub-study. This frailty/random effects model has been recommended for optimizing power in multi-center time-to-event models (26). In addition to intervention versus control arm, another set of “per protocol-like” strata was produced by additionally filtering for FFQ-based self-reported fat reduction (in the intervention group) or fat increase (in the control group). CHD qualitatively showed the expected interaction, i.e. a stronger inverse association between LDL-FRS and disease risk in the fat reduction group). Ischemic stroke showed a similar pattern, with a risk reduction only in the fat reduction group (p = 0.020). In contrast, hemorrhagic stroke, while having a low number of events (44 in total), showed a positive association only in the fat reduction group (p = 0.011). Results for diabetes qualitatively mirrored those for CHD and ischemic stroke, while those for non-CVD death did not vary across groups. These cross-arm differences were generally strengthened when comparing the per protocol-like strata, with a much stronger effect for CHD in the confirmed fat reduction stratum (p = 4.07e-5). In DM trial arm interaction models (score x arm), only ischemic stroke reached nominal statistical significance (p < 0.05).

**Figure 2:**
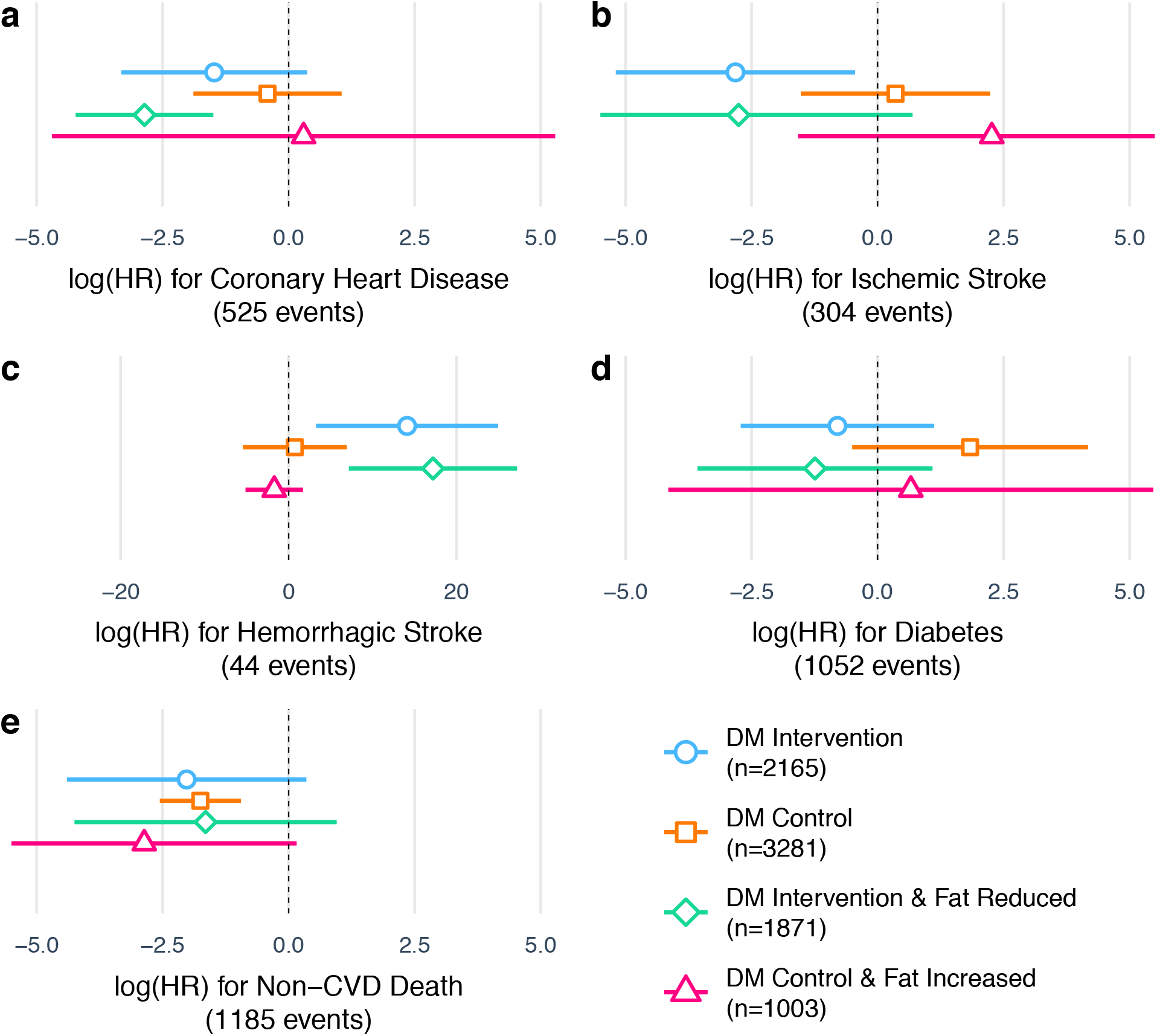
LDL-FRS prediction of chronic disease development. X-axis shows log-hazard ratio estimates for the LDL-FRS from Cox proportional hazards regression for a) coronary heart disease, b) ischemic stroke c) hemorrhagic stroke, d) diabetes, and e) non-CVD death. Separate estimates are shown for DM trial intervention arm, control arm, and the same strata filtered for FFQ-reported fat reduction or increase, respectively. Cox models are adjusted for age at baseline and include a random effect for WHI genotyping sub-study. Error bars represent 95% confidence interval estimates for the regression coefficients.

## Discussion

Diet response scores have shown some success in predicting the response of cardiovascular risk factors (CRFs) to nutritional interventions, but they are often based solely on main effects or single GDI SNPs. Here, we explored the potential for gene diet interaction (GDI)-based diet responder score development, leveraging the multi-trial setup of the Women’s Health Initiative. We developed what to our knowledge is the first example of a diet response score based on a hypothesis-free genome scan for each of six risk factors, and showed preliminary evidence for the viability of a LDL-C fat response score. The set of SNPs used for each score were limited to those showing nominal main effects in large-scale GWAS as a compromise between discovery and utilization of prior information, which was supported by the weaker results in sensitivity models incorporating either stronger (suggestive main-effect) or weaker (all SNPs) variant filters (Supp. Table S2).

Though FRS for six CRFs were developed and tested, only that for LDL-C showed nominal significance in predicting 1-year changes in the corresponding CRF. Multiple factors could explain this lack of predictive performance in general. First, analysis in observational datasets carries with it the potential for confounding of the observed relationships. Second, FFQs are imprecise instruments for measuring dietary intake, and though the FFQs used in the present study were optimized for detection of dietary fat, there was potential for substantial misclassification of this environmental exposure. Third, power calculations shown here and elsewhere suggest that a cohort of this size may not be powered to detect many very small gene-environment interactions such as are sought in genome-wide approaches like the one used here.

*CSMD1, PTPRD*, and *RGS12* stood out as genes containing the highest number of SNPs in the LDL-FRS (11, 9, and 9, respectively, after LD-pruning for r^2^ < 0.5. *CSMD1* variants are notably associated with LDL-C response to statin treatment (27) as well as SBP response to a high-salt diet [Newton-Cheh2009]. *CSMD1* has also shown epigenetic associations with LDL-C (28) as well as response to modification of dietary fat composition (29). *PTPRD* variants modulate the response of T2D patients to pioglitazone therapy (30) and show suggestive associations with eating behaviors (caloric intake at dinner) (31). *RGS12* has been linked to LDL-C in GWAS (32). Altogether, these genes have literature evidence for relationships to dietary intake, response to cardiometabolic therapies, and LDL-C, but have not until now been shown to directly modify the LDL-C response to dietary fat proportions. We note that there is a bias towards identifying LDL-C-related variants in the LDL-FRS, as only nominally-associated main-effect SNPs were used as input to the score development algorithm.

A reasonable body of literature exists establishing GDIs for both dietary fat on CRFs (33,34) and general dietary exposures on LDL-C (35). Multiple studies have looked specifically at genetic variants modulating the LDL-C response to dietary fat. For example, a caloric restriction intervention in type 2 diabetics was more effective in reducing LDL-C in ApoE4 carriers (−15.6% versus −0.7%) (36). Two studies using the POUNDS LOST trial found relationships of specific polymorphisms with 2-year changes in response to a dietary fat intervention. Carriers of a risk allele at the APOA5 variant rs964184 show a decrease of 7.5 mg/dL in LDL-C in low-fat but not high-fat interventions (37), and carriers of the minor allele at CETP rs3764261 show an 8.9 mg/dL greater decrease in LDL-C on a low-fat diet (38). Our observed effect size of a 5.4 mg/dL decrease in LDL-C is of a similar magnitude to these findings, and emerged despite the multi-factorial nature of the WHI DM trial (which incorporated additional nutrition recommendations). The observed variance explained of 3.4% for the LDL-FRS means that the score, while contributing meaningfully to the prediction, does not capture most of the interindividual variability in LDL-C response to the WHI DM trial intervention. Based on prior observations of an inflection point in the impact of various genetic risk scores near the 90th percentile (39), we additionally evaluated the impact of LDL-FRS in the top 10%, finding almost double the LDL-C reduction in DM intervention participants with values at this extreme.

There has been interest in the past in using main-effect genetic risk scores (GRS) as genetic variables in order to improve statistical power to detect gene-environment interactions (40). Such interactions may be viewed from a lens in which genetic risk corresponds to a predisposition that is only triggered in certain environments (e.g. dietary behaviors). Here, we observed only a minor association of a main-effect GRS for LDL-C with greater LDL-C reductions in the DM trial (p=4.5e-27). This trend runs counter to a prior observation of greater lifestyle intervention effectiveness for LDL-C reduction in those with low genetic risk of hyperlipidemia (41). This discrepancy may be due to differences between the DM trial and the personalized diet and lifestyle changes recommended in the intervention in question. Regardless, the meaningful increase in predictive power of the GDI-based FRS compared to the main-effect GRS for LDL-C indicates the value in using interaction-based scores rather than simple genetic predispositions for the development of personalized dietary recommendations.

A diet response score such as that developed here is most useful if its value extends beyond just risk factor changes and predicts downstream changes in chronic disease and mortality risk. Suggestive interactions for CHD, ischemic stroke, and diabetes were apparent across strata (Figure 2a,b,d), corresponding to a decreased risk in fat-reduction participants (whose LDL-C would be expected to drop more prominently according to the score). In contrast, hemorrhagic stroke showed the opposite trend, with a positive score-disease relationship only in the fat reduction group. This result is in line with existing evidence for the detrimental effects of low LDL-C on hemorrhagic stroke risk (42). Non-CVD death showed no major associations, which could be expected due to the dominance of this category by cancer outcomes and the equivocal associations of cancer with lipids (43). We note that all disease outcome relationships assessed here are subject to the major caveat that dietary evolution and decreased adherence likely developed over time in many subjects, diluting the utility of the randomization and 1-year changes used for stratification in Figure 2.

The present study had the advantage of developing a GDI-based dietary fat score in almost 10,000 women and testing in a dietary intervention trial using independent individuals from the same population. However, nominal main-effect SNPs were prioritized to improve statistical power given this moderate sample size, an approach which may fail to identify interactions with effect directions opposite that of the main effect. Smaller fractions of alternate ancestries in this population also made development of ancestry-specific response scores unrealistic. Additionally, the DM trial intervention in which the scores were tested may not exactly match the intervention relevant to the purely fat reduction-focused score developed here; it included additional non-fat-related dietary recommendations that may have effected the interactions examined here, and did not ultimately achieve its intended 20% fat reduction. Finally, this study only examined women, despite the fact that CRF profiles and their genetic trait architectures are known to vary across sexes (44).

In summary, we present a method for the development of diet response scores based on genome-wide gene-diet interaction study summary statistics. While the resulting dietary fat response scores are not all informative, a score focused on LDL-C is predictive of 1-year LDL-C changes during a fat reduction trial. Its performance is risk factor-specific, and is superior to that of a main-effect genetic risk score for LDL-C. Furthermore, it displays suggestive relationships with chronic disease outcomes, following known coronary heart disease biology and adding a new perspective to known discrepancies between associations of LDL-C with cardiovascular disease and stroke subtypes. These findings support the utility of gene-diet interactions for personalized nutrition while highlighting the need for increased sample sizes and improved diet measures for the discovery of robust genetic predictors of diet response.

## Data Availability

Raw data (genomic and phenotypic) are available from dbGaP (details provided in the manuscript).

